# Investigating the potential for machine learning prediction of patient outcomes: a retrospective study of hospital acquired pressure injuries

**DOI:** 10.1101/2020.03.29.20047084

**Authors:** Joshua J. Levy, Jorge F. Lima, Megan W. Miller, Gary L. Freed, A. James O’Malley, Rebecca T. Emeny

## Abstract

**Background:** While recent research efforts to reduce pressure ulcers in the clinical context have focused on key retrospective characteristics, little work has focused on creating real-time predictive models to prevent this avoidable hospital-acquired injury. Furthermore, existing machine learning heuristics often fail to surpass traditional statistical models or provide individual-level risk assessments with explanations for each patient. Thus, we sought to compare the predictive performance of five machine learning and traditional statistical modeling techniques to predict the occurrence of Hospital Acquired Pressure Injuries (HAPI).

**Methods:** Electronic Medical Record (EMR) information was collected from 57,227 hospitalizations, containing 241 positive HAPI cases, acquired from Dartmouth Hitchcock Medical Center from April 2011 to December 2016. The five classifiers were trained to predict HAPI incidence and performance was assessed using the C-statistic or Area Under the Receiver Operating Curve (AUC).

**Results:** Logistic Regression was the best modeling approach (AUC=0.91±0.034). We report discordance between predictors deemed important by the machine learning models compared to traditional statistical model. We provide means to visually assess factors important to every patient’s prediction, regardless of the modeling approach, through Shapley Additive Explanations.

**Conclusions:** Machine learning models will continue to inform decision making processes but should be compared to traditional modeling approaches to ensure proper utilization. Disagreements between important predictors found by traditional and machine learning modeling approaches can potentially confuse clinicians and as such need to be reconciled. Future efforts to analyze time-stamped, prospective medical record data will be enhanced by patient-specific details. These developments represent important steps forward in developing real-time predictive models that can be integrated and readily deployed in electronic medical record systems to reduce unnecessary harm.

## Background

Hospital Acquired Pressure Injuries (HAPI) are preventable medical errors with costly implications for patients, health care institutions and consumers [1]. These injuries arise from a sustained period of compression between a bony surface and an external surface, often due to immobility and shear[2]. The development and occurrence of these events are difficult to detect and localize during early stages due to little superficial presentation and thus provide further motivation for the development of methods that are able to detect and preempt occurrence of HAPIs[3].

Reported rates of HAPIs vary considerably across the United States, which is largely attributed to inappropriate coding and underreporting. Despite the inability to precisely pinpoint the burden of this condition, a prior study from 2012 has indicated that HAPIs have cost the US healthcare system an estimated 6 to 15 billion dollars per year[4]. Most of these costs have been shifted to the hospitals, but patients bear additional liability when factoring for deductibles, co-payments and coinsurance and the additional length of stay needed to treat this condition[5].

Thus, these individual and societal burdens may be reduced by better understanding patient-specific factors associated with HAPI and by using information regularly collected in electronic medical records to develop predictive risk models for prevention of HAPIs. The ability of prediction models to fit a set of data can be evaluated and compared by taking note of the concordance index, otherwise known as the C-statistic or alternatively the area under the receiver operating curve (AUROC/AUC). The receiver operating curve explores changes in the model’s sensitivity and specificity as the predictive threshold for assignment to the positive class (or outcome, i.e. a HAPI event) is changed[6]. The AUC of the fitted model estimates the probability that a randomly selected hospital encounter that resulted in a HAPI event has a greater predictive probability than a randomly selected hospital encounter without a HAPI event. The larger the C-statistic, the better a model is at predicting these adverse events.

A well-known clinical predictor of HAPIs is the Braden Scale, a measure that incorporates information from six sub-scales (sensory perception, moisture, activity, mobility, nutrition, and friction/shear) to arrive at a risk score between 6 to 23, where scores below 9 indicate severe risk [7]. Prior studies that utilized this scoring system yielded C-statistics of 0.67 and 0.77[8, 9]. Nevertheless, the reported low specificity of the measure begs the inclusion of other important predictors. This has led to the expansion and critical evaluation of the covariates sought to predict HAPI incidence[8].

Machine learning, the specification of a model after a heuristic search for the ideal set of non-linear interactions between predictors, may be a useful tool that can enhance clinical encounters for the prediction and reduction of patient risk [10]. Recently, some of these HAPI predictors have been incorporated into logistic regression and machine learning approaches. A 2015 article applied six diverse machine learning algorithms to a cohort of 7,717 ICU patients and reported a C-statistic of 0.83[11], while another study reported a C-statistic of 0.84 for a general hospital population of 8,286 observations using logistic regression with under-sampling of the negative cases during model fitting[12]. Other studies have applied Bayesian Network approaches to Braden subscales[13], random forest[14] and models built off of Electronic Medical Records (EMR) and claims data [15, 16].

Many of these studies attempt to utilize sophisticated machine learning models without critically evaluating whether it is a more appropriate model than traditional statistical techniques that are more readily adoptable by clinicians. Some studies do not include a traditional statistical model baseline [14], while others appear to neglect the implications of the failure to outperform these traditional techniques [11]. In some cases, inappropriate predictors (e.g. those that occur or that are measured in the future) have been included in machine learning models by implementers focused on predictors to such a degree that they bypass questioning whether their model makes sense. In addition to this, none of these models offer/provide intuitive explanations for predicted risk scores for individual patients, but rather report how the important variables behave globally; individual-level information could better inform the clinician’s treatment of a specific patient, thereby reducing these costly medical errors.

We wanted to apply machine learning techniques to one example of patient outcomes, pressure injury prevention, to demonstrate the utilities of such approaches and illustrate the importance of individual-level model explanations. Here, we improve on previous analytical benchmarks through the rigorous evaluation of a diverse set of machine learning and traditional statistical methods. We arrive at a prediction model that can be understood clearly at the individual level and explains the heterogeneity in the patient population to serve as grounds for the development of future personalized real-time predictive models. Finally, based on our results, we critically assess the role of machine learning for the development of retrospective HAPI prediction models. Nonetheless, these applications may augment standard modeling approaches when evaluating real-time prospective data captured through the EMR.

## Methods

### Data Collection, Variable Selection and Preprocessing

The data utilized for our predictive models were acquired from a prior retrospective study conducted at Dartmouth Hitchcock Medical Center from April 2011 to December 2016[8] after approval of an Institutional Review Board. Data was collected from EMR for patients who were 18 years or older; each observation represented an individual’s hospital stay of three or more days and at least 3 recorded Braden scale measurements. This constituted a dataset of 57,227 hospitalizations, containing only 241 positive HAPI cases, which epitomizes a highly imbalanced dataset that requires further techniques to manage such class imbalance.

EMR variables were selected for our study based on prior literature, expert opinion and based off of selection criteria from a previous study [8] (Additional Figure 1, Supplementary Table 1). All individual predictors demonstrated statistically significant associations with HAPIs (Additional Figure 1, Supplementary Table 2), save for ambulatory status and race. We recapitulated the results from Miller et. al. [8] to validate our variable selection; however, we removed the length of stay (LOS) variable because it is not valid for use in a task of predicting an outcome from an interim point of a patient’s stay. We imputed two variables with missing data (Additional Figure 1, Supplementary Figure 1); time in operating room (OR) was imputed with zeros under the assumption that a non-record was never present in the OR, and body mass index was imputed using Multiple Imputation by Chained Equations (MICE)[17]. The data was split into 80% training to update the model parameters and 20% testing for analysis of the ability of the model to generalize to an unseen population. A detailed explanation of the selected variables is included in Additional File 1.

**Figure 1:**
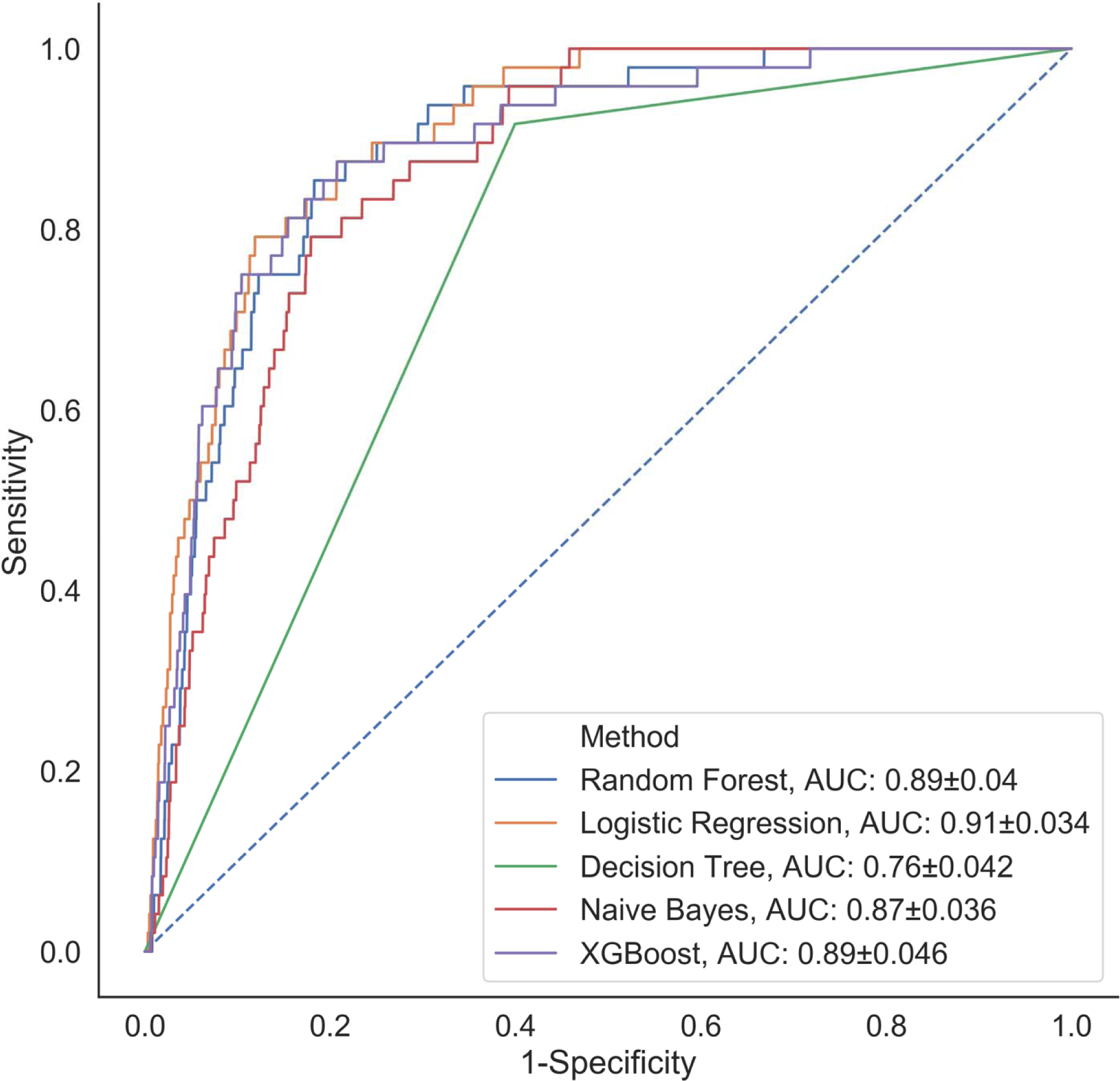
Comparison of classification performance of the five analytical models.

### Description of Modeling Approaches

We performed rigorous evaluations of five different predictive modeling approaches: Naïve Bayes, Decision Trees, Random Forest, XgBoost, and Logistic Regression. We estimated the ideal set of model tuning heuristics for the Decision Tree, Random Forest and XgBoost approaches using an exhaustive grid search on the training set with 5-fold cross-validation. Then, we trained the final predictive model on the training set for all five approaches. The primary metric to assess model performance across a wide range of sensitivity thresholds was the area under the receiver operating curve (AUC). We have included a discussion of each of these analytical techniques in Additional File 1. [18] [19] [20, 21] [22] [23, 24]

### Circumventing Class Imbalance Issues

As aforementioned, there are only 241 HAPI-positive samples in a dataset of 57,227 samples. In accordance with a recent pressure ulcer modeling study that found undersampling negative samples to be an effective modeling technique[12], we employed techniques to upweight the importance of the positive samples [25]. For logistic regression, this meant assigning a higher weight attributed to the positive class, while for random forest techniques, this meant under-sampling the occurrence of negative controls during training time. Experiments with other class balancing techniques such as oversampling and SMOTE (Synthetic Minority Over-Sampling Technique, which over-samples the minority class) [26] appeared to not be as effective as reweighting the model objective and under-sampling during training. Preliminary testing of this technique demonstrated that adding the class balanced weighting marginally improved the AUC of the resultant model. Logistic regression techniques are well-equipped to handle rare events and thus do not usually require class balancing. However, other machine learning models may not explicitly account for rare events and thus require class balancing. To this end, we implemented these class balancing techniques for all models to offer a fair comparison.

### Developing Individual Level Explanations

Many “black box” machine learning models have difficulties in explaining exactly how they arrived at their predictions. The ability to explain predictions in real world applications is paramount to the actual use and applications for HAPI predictions. While a number of explainability techniques seek to find important predictors across all patients as a way to demonstrate how the model is learning, very few methodologies have been developed to explain for each patient what variables the model had found important. Here, we utilized Shapley Additive Explanations (SHAP) [27] to directly indicate the contribution of each predictor to the predicted probability of being associated with a HAPI event. SHAP estimates a linear model for each held-out observation under scrutiny, where the importance of each predictor is given by the unique model coefficients. However, these personalized models, when summing their coefficients across the cohort, are able to find the overall importance of each predictor. While the SHAP importance from a linear modelling approach should exhibit properties of the linear model, SHAP scores for machine learning models indicate variables that are important and specific to each patient. Plots that summarize the behavior of the model predictors over the entire dataset could offer an insightful tool for aiding the clinician to quickly interpret patient symptoms and intervene to prevent HAPI from occurring.

## Code Availability

The results were derived using a custom data pipeline that utilized Jupyter Notebook version 5.7.8 with a Python 3.7.3 Kernel. The model graphics were generated using the SHAP library. We tested for possible interaction effects using the *InteractionTransformer* package [28]. Code is available upon request.

## Results

We fit the five modeling approaches to our HAPI dataset and derived C-statistics on the held-out test set (Figure 1). Out of all of the models, decision trees performed the worst with a C-statistic of 0.76, followed by Naïve Bayes with an AUC of 0.87. Results indicate that the logistic regression model (AUC=0.91) outperforms the other modeling approaches (Random Forest, XGBoost AUC=0.89). These results provide supporting evidence that the logistic regression model identifies the model specification closest to the underlying true model.

Concerns about the transparency of machine learning techniques have been raised by researchers and professionals working in highly regulated environments such as in the practice of law and medicine[29]. While high predictive accuracy is important, understanding how an algorithm makes a recommendation is fundamental to establish trust and foster acceptance. We applied the SHAP methodology to find the overall important global variables that were important for the prediction of the logistic regression, XGBoost and Random Forest models. While we found a significantly strong positive correlation between the importance of the predictors across all three models (Additional Figure 1, Supplementary Tables 3-5), we noted important disagreements between predictors identified by each model with regards to their level of importance. For instance, low nutrition, average activity and moisture were found to be highly important by the Logistic Regression model, but not by the Random Forest or XGBoost models. Alternatively, smoking was upweighted by the Random Forest and XGBoost models, but not by Logistic Regression. All models found low friction, average mobility and whether the patient’s diet was taken by mouth (NPO status) to be important.

While ranking of important predictors can be found in the SHAP summary plots (Figure 2), one useful feature of SHAP, irrespective of modeling approach, is to portray the important predictors that influence the prediction of a single patient. To more closely interrogate the predictive model for individual patients, we assessed a few select force plots (Figure 3) that depict each model’s prediction and the predictors’ importance across select individuals. The logistic regression, random forest and XGboost models all appear to make similar predictions and find similar features to be important for the two observations chosen for display. We have included a figure that showcases the use of this to capture important predictors across 300 patients out of the entire study population (Figure 4). This figure is a static representation of a web-based application that the physician or end-user can interact with to reveal the important predictors for each patient.

**Figure 2:**
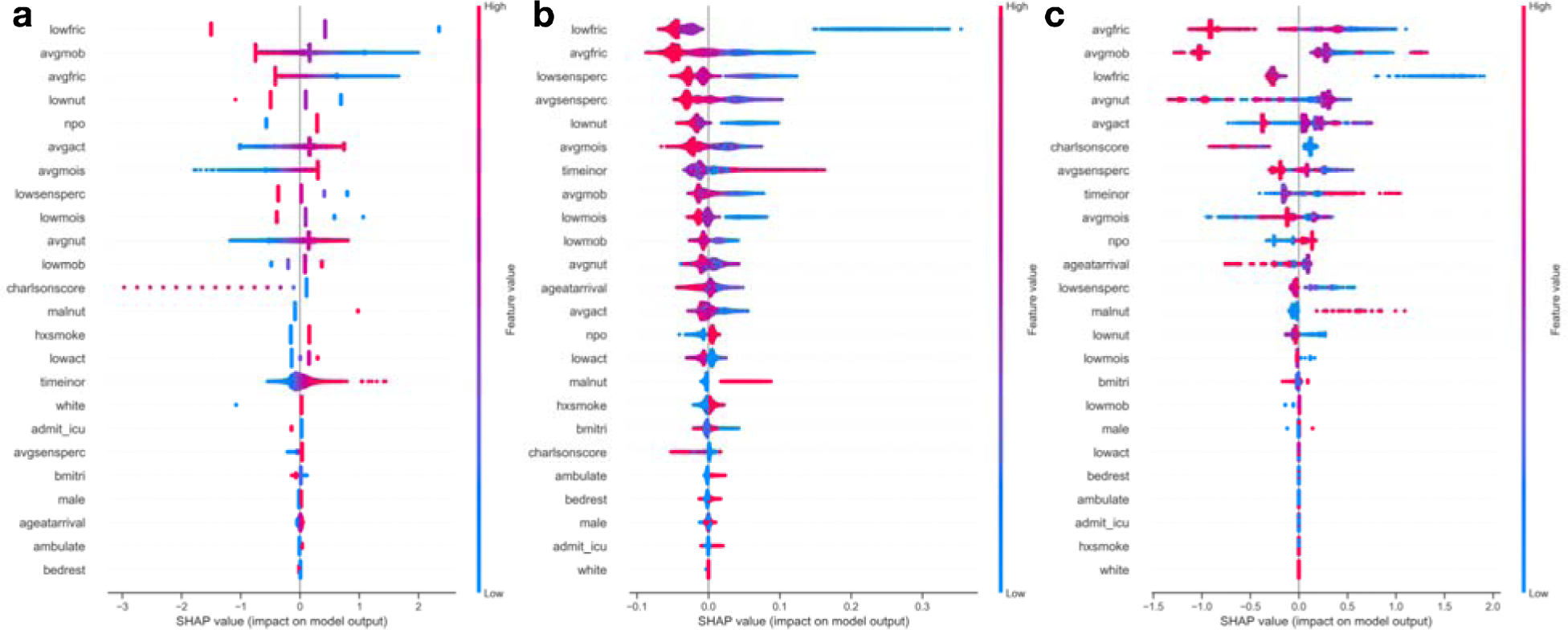
Global predictor importance (SHAP summary plots) of patient specific factors for: a) Logistic Regression, b) Random Forest, c) XGBoost; The plots for each model (a-c) consist of a point per patient hospitalization across all predictors. The points are colored by the features value and lateral displacement from the centerline indicates the importance of that feature for that particular individual. Values that increase the probability of being classified as a HAPI are displayed to the right of the centerline of each plot; red dots indicate a high feature value, while blue dots indicate a low feature value. For instance, increased HAPI incidence was associated with decreases in the Braden subscale score for low friction, average mobility, average friction and low nutrition in the logistic regression plot (a).

**Figure 3:**
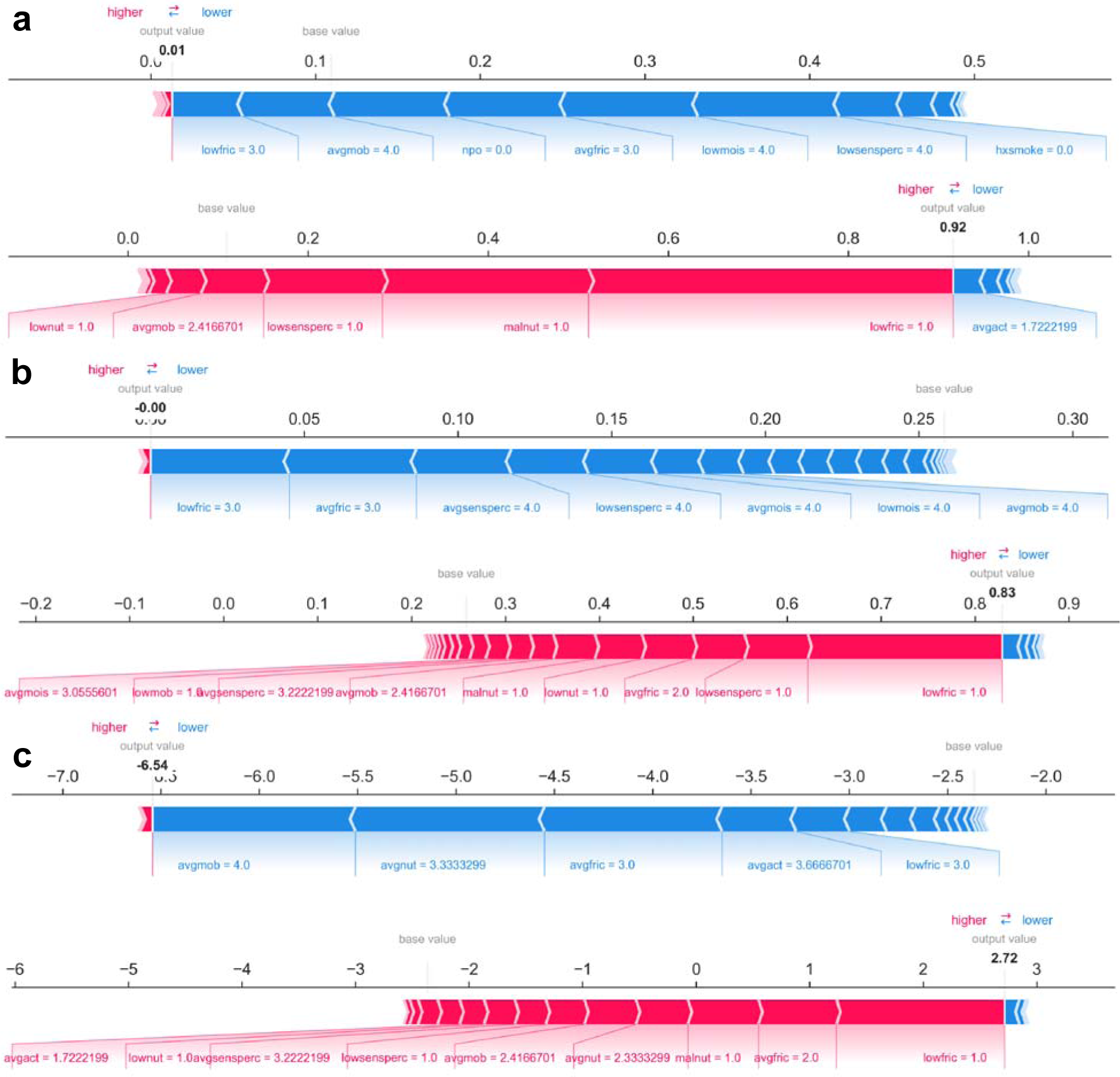
Predictions and decomposition of predictor importance (force plots) for two individuals (top versus bottom of each panel) using: a) Logistic Regression, b) Random Forest, c) XGBoost. The predictors are associated with both increased and decreased HAPI. Certain values (e.g., increasing values) may be associated with one or the other. Blue colors indicate predictors that are associated with decreased HAPI incidence, while red colors indicate predictors associated with increased HAPI incidence; magnitude of each arrow indicates the level of importance of predictor for that prediction

**Figure 4:**
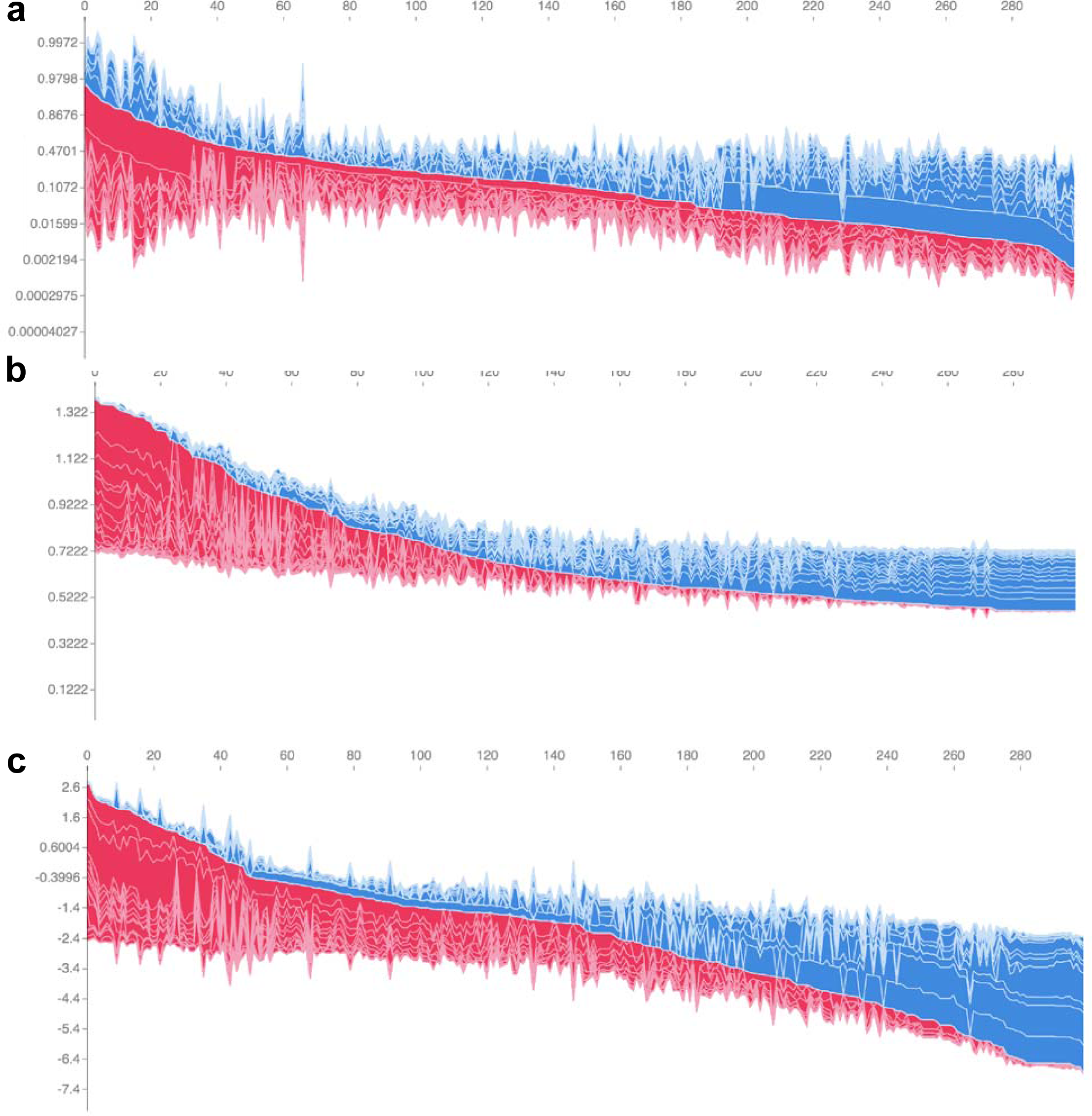
Individual-patient explanations (force-plots) are rotated to a vertical position and stacked horizontally to form interactive plots detailing explanations of HAPI predictions across a large patient population while still allowing interrogation of each patient. We note here that this feature is a web-based interactive plot; the physician or end-user can hover over individuals with their computer mouse, from which the application will display/highlight the important predictors for those individuals. SHAP derived force plots depicting individual predictions and explanations for the first 300 hospitalizations in the study population, ordered from highest HAPI predicted probability (red) to lowest (blue) for: a) Logistic Regression, b) Random Forest, c) XGBoost

Averaging the absolute value of the SHAP scores for each predictor across the cohort derives an overall importance ranking of the predictors. We found that averaging the SHAP importances for the logistic regression model yields an approximation of the standardized regression coefficients (Pearson-r=0.914, p=4.5e-10, average absolute difference=0.08) (Additional Figure 1, Supplementary Tables 6-7, Supplementary Figure 2). This convergence reinforces the notion of correspondence between the totaled SHAP coefficients across all of the individuals and the effect estimates of the Logistic Regression model.

## Discussion

Our study sought to compare the predictive performance of ML and traditional statistical modeling techniques using the example of hospital acquired pressure injuries Thus, we built a predictive risk model for hospital pressure injuries based on a retrospective cohort of over 57,000 hospitalizations over a 5-year study period. Our results indicate that the logistic regression technique outperformed the five other machine learning approaches when applied to retrospective data without temporal changes in patient status. This ideal model specification (0.91 C-statistic) exceeded the performance recorded in prior publications (0.84 C-statistic) and presents opportunities for early detection of symptoms while minimizing the burden on the clinical staff.

However, the fact that logistic regression was able to achieve such remarkable performance indicates that the use of machine learning for HAPI prediction is not optimal given the utilized variables and available retrospective data. This conclusion is not surprising because predictors that vary linearly and continuously with the outcome are better approximated by a line, not the step-function form that tree-based classification algorithms, optimized in machine learning, support. In this context, the selection of the features by expert opinion and testing univariable associations with HAPI outcomes may have biased the selection of our variables to those that are less likely to interact or vary nonlinearly with HAPI risk.

Previous studies have reported the training and utilization of machine learning models without consulting traditional statistical approaches[14]. We find the allure of and immediate acceptance of automated machine learning approaches a cause for concern due to the implications of how it arrives at its decision. From our study, we reported discordance between some of the predictors found important by the Logistic Regression and machine learning-based modeling approaches. This disagreement may potentially confuse the clinician as to which model-learned factors to focus on. The clinician may focus on records of low friction, average mobility, and NPO status if utilizing either the machine learning or Logistic Regression modeling approaches. However, they may choose to more often disregard indicators of low nutrition, activity and high moisture while prioritizing smoking status if opting to utilize the machine learning models over Logistic Regression. Shifting the physician’s attention to these machine learning derived predictors may have unintended consequences for the patient, thus it is imperative to resolve any uncertainty introduced by these machine learning techniques before seeking to adopt them.

In concert with cautionary advice on machine learning implementations, Logistic Regression approaches are more intuitive and easier to understand and currently are more readily adoptable in the biomedical community. The results corroborate with existing literature suggesting that machine learning models are frequently unable to outperform logistic regression models in the clinical setting, although a few other studies have disputed this claim [30–32]. The machine learning models in this study disregarded important predictors, such as nutrition and activity, corroborated by evidence from prior studies, and since these models underperform compared to traditional statistical modeling, it would be a safe option to continue to use the Logistic Regression approach. Nevertheless, in light of recent studies indicating relationships between excluded biomarkers such as albumin and C-reactive protein levels (CRP) [33] in the pressure ulcer setting, having time-stamped data with access to complete biomarker data may warrant us to revisit our modeling approach to incorporate the agility of machine learning techniques in order to specify and explore interactions.

While SHAP coefficients for the Logistic Regression model converge on the global Logistic Regression model coefficients, they provide a quick and intuitive means for obtaining the patient’s risk and how certain predictors contribute to that risk. We further highlight a key difference between SHAP model coefficients and the Logistic Regression coefficients: the logistic regression model beta coefficients are a global descriptor for predictors from the training set, while the SHAP models are fit on the held-out test set and can converge to these coefficients. SHAP is useful for generating explanations for a machine learning model to capture the heterogeneity in the population by fitting separate models for each individual. While SHAP may be less useful for generating interpretations for the linear model, the software offered to produce these patient-level explanations can be easily deployed into an EMR system for clinical use.

There are a few limitations to our study. The study data was collected from a single institution and our patient demographic (97% white) does not correspond to that across the United States. Also, we are unaware of the effect that Dartmouth Hitchcock specific HAPI intervention programs may serve to bias HAPI results [1]. Thus, our results may not generalize to other institutions. It is beyond the scope of this work to explore HAPI predictions outside the hospital setting; although a significant number of pressure injuries occur in long term care facilities, we should be careful to extend conclusions to those patients.

In addition, we were unable to capture all possible clinical covariates or fully utilize real-time repeated measures for this study. The mean length of stay (LOS) for a patient in our study population who does not experience a HAPI is 8.2 days (σ == 9.7) and for those who do experience a HAPI is 30.6 days (a == 28.6). A short length of stay for a HAPI patient may make it difficult to collect enough repeated measurements (at least 3) to make real-time predictions. Since primary pressure ulcers are often overlooked, a reduced observation time may limit our ability to make substantial inferences based on sparse information. A real-time predictive model should account for the impact that the length of stay can have on pressure injury incidence while avoiding issues associated with record completeness. Nevertheless, the addition of repeated lab measurements, unstructured clinical note data, and modalities such as biomedical imaging and sensor data from wearable technology [34–36], would be advantageous towards developing more sophisticated and actionable real-time predictive models.

The use of Shapley feature attributions presents a great opportunity to develop a set of explanatory tools to more quickly assess machine learning predictions for any patient outcome. In this study, we used them as a means of comparison to understand which predictors were found to be important for each machine learning model in predicting pressure injuries. The preliminary inspection of these SHAP scores alerted us to the possibility that the machine learning approaches could potentially mislead the clinician in their treatment of symptoms associated with the occurrence of pressure ulcer injuries. While the ultimate utility in using SHAP lies in the ability to fit explanatory models for each individual in the case that machine learning approaches dominate, SHAP, in any model application, can generate instance-wise importance values for useful, patient-specific readouts for the clinician.

## Conclusions

Machine learning will likely continue to be incorporated into the clinic and inform clinical decision making. Its crescent popularity can be attributed to the promises of better handling large, unstructured, and heterogeneous datasets. We sought to understand how to best utilize these machine learning approaches through extension of its application to pressure injury prevention. In this study, we demonstrated that a Logistic Regression modeling approach outperformed four other machine learning methods for HAPI prediction while improving on existing HAPI prediction benchmarks. In addition, we highlight the potential to integrate patient-level explanations into existing EMR systems. We believe that future applications of machine learning algorithms that exploit repeated measurements, laboratory markers and unstructured clinical notes can provide a promising opportunity to build real-time prediction mechanisms that can be readily embedded into an EMR system to alert clinical staff to high risk patients.

## Data Availability

The EMR dataset curated from Dartmouth-Hitchcock records contains information that could compromise research participant privacy/consent and thus cannot be released due to HIPAA regulations. An IRB approval is required for on-site access and review of the data.

## List of Abbreviations

HAPI: Hospital Acquired Pressure Injuries
EMR: Electronic Medical Records
AUC: Area Under the Receiver Operating Curve; C-Statistic
LOS: Length of Stay
OR: Time in Operating Room
MICE: Multiple Imputation by Chained Equations
SMOTE: Synthetic Minority Over-Sampling Technique
CRP: C-Reactive Protein Levels
NPO: Patient’s Diet Taken by Mouth
SHAP: Shapley Additive Explanations

## Ethics Approval and Consent to Participate

All relevant ethical guidelines have been followed and any necessary IRB and/or ethics committee approvals have been obtained. All necessary patient/participant consent has been obtained and the appropriate institutional forms have been archived.

## Consent for Publication

Not Applicable.

## Competing Interests

The authors declare that they have no financial or non-financial competing interests.

## Funding

The Dartmouth Clinical and Translational Science Institute supported RTE under the award number UL1TR001086 from the National Center for Advancing Translational Sciences (NCATS) of the National Institutes of Health (NIH). JJL is supported by the Burroughs Wellcome Fund Big Data in the Life Sciences training grant at Dartmouth. The funding bodies above did not have any role in the study design, data collection, analysis and interpretation, or writing of the manuscript.

## Authors’ Contributions

All authors were responsible for the study design, data collection and statistical analysis, writing of the manuscript and decision for the manuscript’s submission.

## Acknowledgements

Not Applicable

## Notes

### Competing Interest Statement

The authors have declared no competing interest.

